# Neutrophil levels correlate with quantitative extent and progression of fibrosis in IPF: results of a single-centre cohort study

**DOI:** 10.1101/2023.09.05.23295078

**Authors:** Andrew Achaiah, Emily Fraser, Peter Saunders, Rachel Hoyles, Rachel Benamore, Ling-Pei Ho

## Abstract

**Background:** Idiopathic pulmonary fibrosis (IPF) is a progressive fibrotic lung disease with poor prognosis. Clinical studies have demonstrated association between different blood leukocytes and mortality and FVC decline. Here we question which blood leukocyte levels are specifically associated with progression of fibrosis, measured by accumulation of fibrosis on CT scan using a standardised automated method.

**Methods:** Using the CALIPER (Computer-Aided Lung Informatics for Pathology Evaluation and Rating) CT algorithm, we determined the correlation between different blood leukocytes (<4 months from CT) and total lung fibrosis (TLF) scores, pulmonary vessel volume (PVV), FVC% and TLCO% at baseline (n=171) and with progression of fibrosis (n=71), the latter using multivariate Cox regression.

**Results:** Neutrophils (but not monocyte or lymphocytes) correlated with extent of lung fibrosis (TLF/litre) (r=0.208, p=0.007), PVV (r=0.259, p=0.001), FVC% (r=-0.127, p=0.029) at baseline. For the 71 cases with repeat CT; median interval between CTs was 25.9 (16.8-39.9) months. Neutrophil but not monocyte levels are associated with increase in TLF/litre [HR 2.66, 95%CI, 1.35-5.25, p=0.005].

**Conclusion:** Our study shows that neutrophil rather than monocyte levels correlated with quantifiable increase in fibrosis on imaging of the lungs in IPF, suggesting its relative greater contribution to progression of fibrosis in IPF.

**Key messages:** *WHAT IS ALREADY KNOWN ON THIS TOPIC:* Idiopathic pulmonary fibrosis (IPF) is a progressive fibrotic condition. Recently, several human studies have implicated blood leukocyte levels (monocyte, neutrophil and lymphocyte) with FVC decline and mortality. However, direct association between leukocytes and progression of fibrosis using quantitative CT analysis has not been explored.

*WHAT THIS STUDY ADDS:* This study explored the association between blood monocytes, neutrophils and lymphocytes against increase in fibrosis over time, measured using a quantitative CT algorithm, CALIPER. We show that levels of blood neutrophil and lymphocytes but not monocytes were associated with greater risk of progression of fibrosis.

*HOW THIS STUDY MIGHT AFFECT RESEARCH, PRACTICE AND/OR POLICY:* Our study shows that neutrophil rather than monocyte levels correlated with quantifiable increase in fibrosis on imaging of the lungs in IPF, suggesting its relative greater contribution to progression of fibrosis in IPF.

## Introduction

Idiopathic pulmonary fibrosis (IPF) is a progressive fibrotic condition. Animal studies have implicated innate immune cells, particularly monocytes and macrophages in the pathogenesis of IPF.^1^ This is supported by several clinical studies which showed that higher blood monocytes levels are associated with mortality in IPF.^2^ Blood monocyte levels also correlated with the extent of fibrosis on CT scan,^3^ and Kreuter also found an association with a composite measure of IPF outcome (FVC decline, 6-minute-walk distance reduction, acute exacerbation and/or mortality).^4^ Moreover, we and others found that the neutrophil:lymphocyte ratio is also a predictor of mortality and FVC decline,^5,6^ and patients with higher levels of neutrophils were more likely to progress from indeterminate for UIP CT pattern to UIP pattern and a clinical diagnosis of IPF.^7^

However, mortality and FVC decline may not accurately reflect progression of fibrosis. Death can be due to other causes e.g. cardiovascular diseases, especially in the elderly population of IPF. Although FVC decline of >10% in IPF is the validated measure of disease progression predictive of mortality,^8^ and serial FVC change the recommended monitoring variable in international guidelines,^9^ this global metric may not accurately reflect regional morphologic changes indicative of progression of fibrosis. In fact a *r*ecent large study suggests that FVC decline in IPF patients is heterogenous and may have little correlation with increase in fibrosis in some patients.^10^

The Computer-Aided Lung Informatics for Pathology Evaluation and Rating algorithm (CALIPER), is an automated quantitative CT application which characterises lung parenchymal features on volumetric high-resolution CT (HRCT), thus quantifying abnormal lung.^11^ In IPF studies it is highly predictive of mortality,^12^ but also enhances risk stratification and cohort enrichment for trial end points.^13,14^ Until now, it has not yet been used to explore leukocyte association with progression of fibrosis.

The aim of the study is two-fold – (i) to assess if leukocyte levels are linked to progression in fibrosis *per se*, and (ii) to explore which leukocytes demonstrate greatest association with fibrosis. We used CALIPER to provide a standardised automated, method of quantifying lung fibrosis.^11^

## Methods

### Study design and patients characteristics

We performed a retrospective analysis of a cohort of patients with IPF, who presented to the Oxford Interstitial Lung Disease (ILD) Service between September 2016 and November 2021. All patients with a multi-disciplinary team (MDT) diagnosis of IPF,^15^ and a CALIPER-compatible HRCT (non-contrast, supine, volumetric HRCT scan) were included.

We asked two main questions – (i) what is the correlation between contemporaneous blood leukocyte levels (within 4 months of the first CT) and baseline amount of lung fibrosis, and (ii) which blood leukocyte levels (neutrophils, lymphocytes, monocytes and their derived ratios) correlated with progression in amount of lung fibrosis. Association between blood leukocytes and FVC decline and mortality were also examined as comparator end points.

### Blood leukocyte measurement

Neutrophil, lymphocyte, and monocyte levels captured from standard clinical ‘full blood count’ analysis within 4 months of initial CT (CT1). Neutrophil:lymphocyte ratio (NLR), monocyte:lymphocyte ratio (MLR) and systemic inflammation response index (SIRI)(neutrophil ×monocyte/lymphocyte) were calculated as described previously.^5^

### CT scans

Non-contrast, supine, volumetric HRCT scan were acquired using a 64-detector row CT scanner. Images were reconstructed using a high spatial resolution algorithm. Non-contrast, volumetric, high resolution CT scans for appropriate subjects were acquired (0.625mm slice thickness at an interval of 0.625mm).

### CALIPER

Full CALIPER data acquisition and processing are described in online Supplemental eMethods. Briefly, classification of parenchymal features was applied to 15×15×15 voxel volumes of interest (VOI) using texture analysis and computer-based algorithmic interpretation of volumetric histogram signature mapping features.^11^ CALIPER evaluation included characterisation and quantification of each VOI into one of five radiological parenchymal categories: normal lung, hyperlucent, ground glass opacity (GGO), reticular opacity and honeycombing. Parenchymal features were expressed as a relative percentage of CALIPER-derived total lung volume. Total lung fibrosis (TLF) represented the sum of GGO, reticular and honeycomb percentages.^14^. We included GGO as these had been reported by our radiologists to be found in areas of reticulation or traction bronchiectasis at MDTs and designated as fine fibrosis in keeping with agreed radiological assessments^16^

Assessment of pulmonary vessel volume (PVV) was also performed prior to pulmonary vessel extraction from lung parenchyma using a multi-scale tubular structure enhancement filter.^17^ This assessment excluded large vessels at the hilum of the lung. The PVV was calculated as absolute volume (cm^3^) and expressed as percentage of total CALIPER-derived lung volume.

The key variables were total lung fibrosis (TLF) [the sum of reticulation, ground glass opacification (GGO) and honeycomb, as % of CALIPER-derived lung volume], TLF/litre (TLF expressed as % of lung volume) and PVV. TLF/litre was used to account for difference in lung volume on serial scans with progression in fibrosis. ^18^ %change in fibrosis is (CT2 TLF/litre – CT1 TLF/litre)/CT1 TLF/litre x100.

### Lung function tests

Baseline spirometry (FEV1 and FVC), transfer Factor of Lung for Carbon Monoxide (TLco), and composite physiological index (CPI)^19^ within 3 months of CT were recorded; all as % predicted for age, height and sex. Annualised change in lung function was calculated as the relative (%) change in absolute value divided by time (years) between tests.

### Statistical analysis

Pearson correlation was used to explore association between blood leukocytes with baseline CALIPER-variables and pulmonary function tests. Cox proportional hazard modelling was employed to test association between each blood leukocyte level (monocytes, lymphocytes and neutrophils) and outcomes of disease progression - (i) ≥7.8% increase in TLF/litre (7.8% being the lower limit of upper tertile) (model A), (ii) ≥ 10% increase in TLF/litre (10% as an arbitrary value) (model B) and (iii) ≥10% FVC decline between CT1 and CT2 (model C). Median and upper quartiles for model A were also tested in preliminary modelling but no significant correlation was found. Harrell’s Concordance index (C-index) was used to assess model strength, describing how well a model can discriminate between two survival distributions.^20^ Kaplan-Meier analysis (Log rank test) was used to evaluate time to all-cause mortality from first CT. A censoring time of 1^st^ January 2022 was applied.

All analyses were performed using GraphPad Prism version 9.2 (La Jolla, California, USA) or SPSS version 27 (IBM Armonk, NY, USA).

### Ethical approval

The study was part of a study examining factors associated with progression of IPF (ethical approval 14/SC/1060 from the Health Research Authority and South-Central National Research Ethics Service).

## Results

### Patients

171 eligible patients were identified. Of these, n=71 had at least one further follow-on HRCT (the CT with the longest interval from the first were selected). Demographics are shown in Table 1. Median interval between CT1 and CT2 was 25.9 (16.8-39.9) months. Median time from first CT to death was 26.2 months (12.4-43.4) vs 45.7 (34.6-61.5) in those surviving to censoring date.

**Table 1.** Baseline characteristics for patients, at the point of first CT used for CALIPER analysis. All values are median (IQR) unless stated. NLR - neutrophil: lymphocyte ratio, MLR - monocyte: lymphocyte ratio, SIRI; systemic inflammation response index, CPI - Composite Physiological Index as calculated by Wells.^19^ 49% of the cohort had ‘probable UIP’ as defined by the 2018 ATS criteria (all had clinical IPF from MDT diagnosis), explaining why some patients do not have honeycomb. Low attenuation areas represent emphysematous areas.

|  | All patients<br>(n=171) | Patients for<br>analysis of<br>progression |
| --- | --- | --- |
| **Demographics** |  |  |
| Age at CT in years | 75.1 (70.1-81.1) | 73.6 (68.3-79.3) |
| Female, n (% of cohort) | 17 (9.9) | 6 (8) |
| Male, n (% of cohort) | 154 (90.1) | 65 (92) |
| **Smoking status** |  |  |
| Never smoker, n (% of cohort) | 47 (27.5) | 21 (29.6) |
| Ex-smoker, n (% of cohort) | 89 (52.3) | 29 (40.8) |
| No data, n (% of cohort) | 35 (20.2) | 21 (29.6) |
| **CALIPER Parenchymal features** |  |  |
| Total lung fibrosis or TLF, as % of lung volume | 14% (8-22) | 13% (7-19) |
| Low attenuation areas, as % of lung volume | 1% (0-2) | 1% (0-3) |
| Total Ground Glass, as % of lung volume | 5% (3-13) | 5% (2-10) |
| Total Reticular, as % of lung volume | 5% (3-9) | 5% (3-8) |
| Total Honeycombing, as % of lung volume | 1% (0-1) | 0% (0-1) |
| **CALIPER Pulmonary vessel volume** |  |  |
| Total PVV in cm<sup>3</sup> | 160 (128-201) | 168 (134-202) |
| Total PVV as % of lung volume | 4.02% (3.02-5.21) | 4.60% (3.04-6.62) |
| **Lung function** |  |  |
| FEV1, litre | 2.19 (1.83-2.68) | 2.34 (1.93-2.83) |
| %FEV1 | 81.2 % (70.9-92.2) | 84.7 % (71.53-91.15) |
| FVC, litre | 2.8 (2.28-3.365) | 2.91 (2.35-3.56) |
| %FVC | 76.57% (66.6-91.15) | 76.44% (68.9-92.37) |
| FEV1: FVC | 80.6 (75.0-86.1) | 82.1 (75.1-88.6) |
| TLCO, SI | 4.56 (3.49-5.48) | 4.9 (3.87-5.80) |
| %TLCO | 56.85% (49.5-67) | 60.05% (51.1-71) |
| CPI Score | 67.48 (61.05-73.12) | 65.36 (59.64-71.25) |
| **Blood leukocytes** |  |  |
| Monocyte x 10<sup>3</sup>/μl | 0.67 (0.58-0.84) | 0.66 (0.55-0.8) |
| Neutrophil x 10<sup>3</sup>/μl | 4.90 (3.85-6.53) | 4.87 (3.85-6.45) |
| Lymphocyte x 10<sup>3</sup>/μl | 1.62 (1.31-2.23) | 1.64 (1.43-2.27) |
| MLR | 0.37 (0.32-0.55) | 0.37 (0.29-0.5) |
| NLR | 2.70 (2.08-4.08) | 2.69 (2.06-3.61) |
| SIRI | 1.79 (1.4-3.08) | 1.76 (1.27-2.63) |
| **Comorbidity** |  |  |
| Reflux, n (% of cohort) | 54 (39.1) | 25 (39.7) |
| PHT, n (% of cohort) | 11 (8.0) | 4 (6.3) |
| IHD, n (% of cohort) | 28 (20.3) | 15 (23.8) |
| Cardiomyopathy, n (% of cohort) | 6 (5.8) | 1 (2) |
| Arrhythmia, n (% of cohort) | 9 (6.5) | 2 (3.2) |
| Hypertension, n (% of cohort) | 29 (21) | 16 (25.4) |
| PE, n (% of cohort) | 3 (2.2) | 2 (3.2) |
| COPD/Emphysema, n (% of cohort) | 8 (7.7) | 4 (8.2) |
| Type II diabetes mellitus, n (% of cohort) | 15 (10.9) | 7 (11.1) |
| **Antifibrotics** |  |  |
| Antifibrotic use, n (% of cohort) | 69 (40.3) | 36 (57.1) |

### Association between blood leukocyte levels and baseline CALIPER and lung function measures

At baseline (CT1, n=171), there was significant correlations between baseline % predicted FVC, TLCO and CPI, and TLF, TLF/litre, and PVV (Table 2). In terms of correlation between leukocytes and CALIPER variables and lung function, there was significant correlation between neutrophil levels and TLF/litre (r=0.208, p=0.007), total PVV (r=0.259, p=0.001), and FVC (r=-0.127,p=0.029) (Table 3). No other leukocyte levels or their derived measures showed significant corelations with Caliper or lung function at baseline (Table 3 and Supplemental eTable S1).

**Table 2.** Association between lung function and CALIPER parameters (Pearson’s correlation) at baseline; n=171 patients.

| CALIPER baseline metrics | FVC% |  | TLCO% |  | CPI |  |
| --- | --- | --- | --- | --- | --- | --- |
|  | r | p | r | p | r | p |
| Lung volume (litre) | 0.634 | **<0.0001** | 0.411 | **<0.0001** | -0.112 | 0.156 |
| TLF (%) | -0.392 | **<0.0001** | -0.461 | **<0.0001** | 0.275 | **<0.0001** |
| TLF (/litre) | -0.430 | **<0.0001** | -0.444 | **<0.0001** | 0.246 | **0.002** |
| Low attenuation areas (% of total | 0.217 | 0.061 | 0.107 | 0.177 | 0.066 | 0.404 |
| GGO (% of total lung volume) | -0.286 | **<0.0001** | -0.316 | **<0.0001** | 0.175 | **0.005** |
| Reticulation (% of total lung volume) | -0.306 | **<0.0001** | -0.407 | **<0.0001** | 0.285 | **<0.0001** |
| Honeycomb (% of total lung volume) | -0.039 | 0.534 | -0.087 | 0.173 | 0.134 | **0.033** |
| PVV (cm<sup>3</sup>) | -0.068 | 0.284 | -0.322 | **<0.0001** | 0.253 | **<0.0001** |
| PVV (%) | -0.448 | **<0.0001** | -0.473 | **<0.0001** | 0.262 | **<0.0001** |

**Table 3.** Correlation of blood leukocytes with CALIPER parameters and lung function (n=171 patients, Pearson’s correlation).

| Blood leukocytes<br>Baseline metrics | Monocytes |  | Neutrophils |  | Lymphocytes |  |
| --- | --- | --- | --- | --- | --- | --- |
|  | r | p | r | p | r | p |
| **CALIPER scores** |  |  |  |  |  |  |
| TLF (/litre) | 0.040 | 0.606 | 0.208 | **0.007** | 0.136 | 0.082 |
| PVV (%) | 0.110 | 0.164 | 0.259 | **0.001** | 0.019 | 0.809 |
| **Lung function tests** |  |  |  |  |  |  |
| FVC% | -0.096 | 0.101 | -0.127 | **0.029** | -0.014 | 0.814 |
| TLCO% | -0.104 | 0.080 | -0.104 | 0.079 | -0.085 | 0.155 |
| CPI | 0.103 | 0.081 | 0.091 | 0.127 | 0.022 | 0.708 |

### Association between blood leukocyte levels and progression of fibrosis

Neutrophil count was significantly higher in cases demonstrating progression of fibrosis as defined by ΔTLF >10%/Ltr [4.87 (3.86-5.66) vs 4.84 (3.85-6.45), p=0.043]. No other differences in leukocyte measure between cases with disease progression and stability was identified. (Supplemental eTable S2).

In model A of Cox proportional hazard analysis, neutrophil levels [HR 1.81, 95%CI 1.10-2.99, p=0.020] were significantly associated with increase in TLF/litre ≥7.8% (Table 4). Lower lymphocyte levels were also significantly associated with TLF/litre ≥7.8% [HR 0.26, 0.08-0.91, p=0.034]. Similar findings were observed in model B (where outcome was TLF/litre ≥10%), except that lower lymphocyte levels were not significantly associated with progression of >10%/Litre. In both models A and B, there was a trend of higher monocyte levels with progression of fibrosis, but this was not statistically significant.

**Table 4.** Cox Proportional hazard analysis for progression of lung fibrosis Hazard ratios in multivariate model generated for outcomes of increase in fibrosis on follow on CT scan in three models as described in three models – Model A - Increase in TLF > 7.8 %/litre, (B) increase in fibrosis>10% /litre and (C) relative decline in absolute FVC >10%. Change in lung volume is measured between CT1 and CT2. Leukocyte levels are presented as continuous variables (outcome for dichotomised values are shown in Supplemental Table S3).

| Multivariate Cox regression | HR | 95%CI | p |
| --- | --- | --- | --- |
| **Model A: Increase in TLF $\geq 7.8\%$ /litre between CT1 and CT2** |  |  |  |
| -Age at CT | 0.96 | 0.88-1.05 | 0.404 |
| -Male | 0.03 | 0.01-0.56 | **0.019** |
| -Change in lung volume (%) | 0.86 | 0.80-0.93 | **<0.001** |
| -Total lung fibrosis (/litre) on 1st CT | 1.12 | 1.05-1.20 | **0.001** |
| -Total PVV (%) | 1.03 | 1.01-1.05 | **0.014** |
| - Low attenuation areas (%) | 1.06 | 0.94-1.20 | 0.318 |
| -Monocyte ($\times 10^3/\mu\text{l}$) | 1.35 | 0.10-18.38 | 0.820 |
| -Neutrophil ($\times 10^3/\mu\text{l}$) | 1.81 | 1.10-2.99 | **0.020** |
| -Lymphocyte ($\times 10^3/\mu\text{l}$) | 0.26 | 0.08-0.90 | **0.034** |
| *Harrell's Index of concordance = 0.91* |  |  |  |
| **Model B: Increase in TLF $\geq 10\%$ /litre between CT1 and CT2** |  |  |  |
| -Age at CT | 0.93 | 0.84-1.04 | 0.208 |
| -Male | 0.02 | 0.02-0.46 | **0.016** |
| -Change in lung volume (%) | 0.82 | 0.73-0.92 | **<0.001** |
| -Total lung fibrosis (/litre) on 1st CT | 1.13 | 1.03-1.25 | **0.013** |
| -Total PVV (%) | 1.04 | 1.01-1.07 | **0.037** |
| -Low attenuation areas (%) | 1.10 | 0.97-1.25 | 0.142 |
| -Monocyte ($\times 10^3/\mu\text{l}$) | 2.37 | 0.09-62.20 | 0.604 |
| -Neutrophil ($\times 10^3/\mu\text{l}$) | 2.66 | 1.35-5.25 | **0.005** |
| -Lymphocyte ($\times 10^3/\mu\text{l}$) | 0.30 | 0.07-1.24 | 0.096 |
| *Harrell's Index of concordance = 0.93* |  |  |  |
| **Model C: Decline in FVC $>10\%$ between CT1 and CT2** |  |  |  |
| -Age at CT | 1.01 | 0.92-1.11 | 0.856 |
| -Male | 0.40 | 0.07-2.31 | 0.308 |
| -%FVC baseline | 0.96 | 0.92-0.99 | **0.049** |
| -Monocyte ($\times 10^3/\mu\text{l}$) | 0.96 | 0.03-28.51 | 0.979 |
| -Neutrophil ($\times 10^3/\mu\text{l}$) | 1.17 | 0.91-1.52 | 0.222 |
| -Lymphocyte ($\times 10^3/\mu\text{l}$) | 0.71 | 0.30-1.71 | 0.448 |
| *Harrell's Index of concordance = 0.90* |  |  |  |

In model C (where outcome was change by 10% of FVC), no leukocyte variable significantly associated with FVC decline. A lower baseline FVC was predictive of FVC decline [HR 0.96, 0.92-0.99, p=0.049].

All leukocyte derived indexes were significantly associated with progression in fibrosis (TLF/litre by 7.8% and 10%) in multivariate analysis (Supplemental eTable S3). 50.7% of patients that underwent repeat CT were receiving antifibrotics at first CT. When adjusted for antifibrotic use, significance was preserved for neutrophils, NLR and SIRI (Supplemental eTable S4).

### Association between blood leukocyte levels and mortality

During the study period, 60 all-cause deaths (35.1%) were reported in this cohort. Leukocyte levels were dichotomised by median values (Figure 1) or normal reference range limits (Supplemental eFigure S1). Significantly shorter survival times were observed for cases dichotomised by median monocyte count (p=0.033) and neutrophil levels (p=0.0180)(Figure 1), and for cases dichotomised by median NLR, MLR and SIRI values (Supplemental eFigure S1). Shorter survival times were also observed for those with TLF, PVV and FVC greater than the median value (TLF >3.30%/L, p<0.001; PVV >4.02%, p<0.001 and FVC >76.6%, p<0.001) (Figure 1).

**Figure 1.**
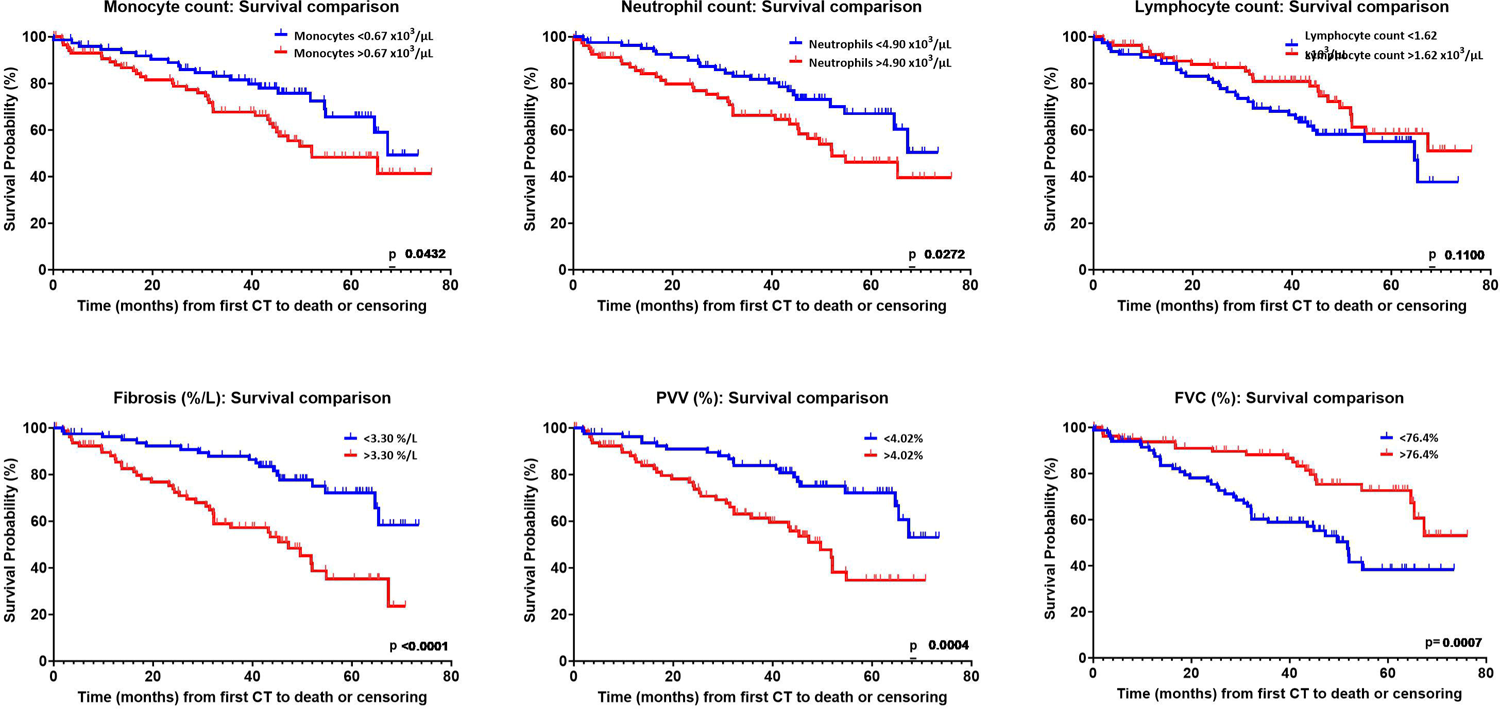
Kaplan-Meier curves for time to mortality for monocyte, neutrophil, lymphocyte levels, TLF, PVV and FVC, all at baseline. Leukocyte levels and CALIPER variables were dichotomised by median value.

## Discussion

In this study, we showed that when progression of disease is categorised specifically by increase in amount of fibrosis, quantified by an automated quantitative scoring modality, neutrophil levels rather than monocytes were the key immune correlate with progression in amount of fibrosis in the lungs. MLR, NLR and SIRI were also associated with progression. Although monocyte count was not associated with disease progression in this cohort, importantly, as with other studies, higher monocyte levels in our cohort were associated with mortality.^4,21^ Our findings suggest that neutrophils may be a contributor to active accumulation of fibrosis in IPF, supported by previous findings of high neutrophils in bronchoalveolar lavage (BAL) of IPF patients,^22^ and more recent findings of higher neutrophil:lymphocyte ratio in IPF patients with greater rates of FVC decline.^5^

Neutrophils have long been associated with immunopathogenesis of IPF. Neutrophilia in bronchoalveolar lavage (BAL) specimens from IPF patients is associated with earlier mortality.^22^ Neutrophil elastases (NE) are elevated in IPF BAL samples,^23^ and experimental data using murine models suggest that NE activates transforming growth factor-ß (TGF-ß) pathway and fibroblast proliferation.^24^ An intriguing and newly identified fibrosis-promoting function of neutrophils is generation of neutrophil extracellular traps (NETs).^25^ These pro-inflammatory collections of chromatin and neutrophils regulate both immune cell function and fibroblast activation.^26^ Whilst a specific association with IPF has yet to be fully described, enhanced detection of intra-pulmonary NETs has been reported in bleomycin models and in non-IPF fibrotic ILD studies.^27,28^ Here, we are able to link blood neutrophil level measured at baseline CT specifically with progression in amount of fibrosis over time. Further prospective translational studies are required to establish the regulatory role of neutrophils, NET formation over time and cytokine and chemokine activity at different stages of fibrosis.

In addition to TLF change, PVV measures also correlated with neutrophil count. PVV is an intriguing quantitative variable that has proved a consistent predictor of disease severity, progression, and mortality in ILD studies.^12,14,29–31^ Our findings are supportive of the original work performed by Jacob et al who demonstrated that PVV% was independently associated with FVC decline,^32^ and Chung et al who demonstrated PVV negatively correlated with TLCO%.^33^ In our study, PVV was independently associated with progression of fibrosis. The pathological mechanisms linking PVV with adverse ILD outcomes are not fully understood and several theories have been suggested. Blood perfusion is reduced in areas of pulmonary fibrosis,^34^ but increased in adjacent areas of unaffected lung.^35^ ln 2017 Jacob et al postulated that correlation between ILD extent and vessel calibre may represent regional elevation in pulmonary artery pressures in mildly fibrotic lung or destruction of the capillary bed in more advanced disease leading to neovascularisation and diversion of blood to unaffected lung areas.^12^ Another possible explanation relates to the negative intrathoracic pressures required of non-compliant fibrotic lungs to generate adequate inspiratory volumes. This could in turn exert additional “tractional” force on the lung vasculature resulting in dilatation in fibrotic regions in a traction-like phenomenon, akin to traction bronchiectasis.^33^

The trend of lower lymphocytes count with progression of fibrosis mirrors the findings of previous studies.^5,36^ Lymphocytic aggregates are a recognised pathologic feature of IPF lesions.^27,37^ The association between low blood lymphocyte count and adverse outcomes in IPF is currently unknown but could be explained in part by lymphocyte dysfunction,^38^ and the sequestering of lymphocytes into sites of inflammation, such as the fibrotic lung. We note that post-hoc analysis of the ASCEND and CAPACITY studies Nathan et al reported that serial increase in NLR over 12 months was associated with mortality.^39^

In our study we report association between higher neutrophil count and lower lymphocyte count taken from full blood count analysis with progression of fibrosis scoring on HRCT. To our knowledge, this specific association has not been reported elsewhere. Other published studies and post-hoc analyses have only reported leukocyte association with mortality, hospitalisation and FVC decline.

Our findings should be framed within the following limitations. Firstly, the retrospective nature of the study meant that we selected patients who had repeat CT scans, which were clinically indicated. This introduced a selection bias towards patients who had a clinical reason to have a CT scan – often, this is worsening in disease either over time or acutely, and CT scans would be performed at different / non-uniform time intervals. Unfortunately, we did not have access to blood leukocyte data measured at the time of the second CT scan. Analysing blood leukocytes / ratios at second CT could may have provided additional information, especially with regards to how any relative change in blood count / ratio may differ between patients with progression of fibrosis or stability at second CT scans. Although the leukocyte levels were measured at the time of the first (presenting) CT scans, this group of patients could have a different disease trajectory compared to those who did not have a second CT scan. There is a possibility that neutrophil levels are more likely a driver in those who are clinically deteriorating. The baseline correlation data (n=171 patients), however, are not limited by these factors. Further analysis of prospective validation cohorts would be required to investigate whether neutrophil count, measured in peripheral blood, could identify patients at greater risk of progression of fibrosis in IPF, and consequently facilitate prognostication of the disease.

From the point of fibrosis scoring, the sum of GGO, reticulation and honeycombing was used as the total lung fibrosis score.^14^ Although each scan was reported by a radiologist who ‘called’ the GGO as fine fibrosis and the overall pattern as probable or definite UIP, other causes such as pulmonary oedema, acute exacerbation and infection could have contributed to this CT finding.^40^

Nevertheless, our study shows that greater neutrophil count was significantly associated with quantifiable increase in fibrosis on imaging of the lungs in IPF. Further studies will be required to validate this findings.

## Data Availability

All data produced in the present study are available upon reasonable request to the authors

## Acknowledgement

The study was funded by the National Institute for Health Research (NIHR) Oxford Biomedical Research Centre (BRC). LPH is supported in part by MRC UK (MC_UU_00008/1).

