## Supplemental for "Neutrophil levels correlate with quantitative extent and progression of fibrosis in IPF: results of a single-centre cohort study"

### **Supplementary materials**

#### **Methods**

##### **CT CALIPER evaluation**

###### **Data processing**

The CALIPER lung texture algorithm (v2.1) was acquired from IMBIO, Minneapolis, USA (www.imbio.com) and installed onto syngo.via software (www.siemens-healthineers.com), a multi-modality reading tool integrated into the local hospital trust system (“client-server platform”). Non-contrast, volumetric, high resolution CT scans (0.625mm slice thickness at an interval of 0.625mm) for appropriate subjects were selected and the Digital imaging and communications in medicine (DICOM) formatted images were uploaded into syngo.via from the local a picture archiving and communications system (PACS).

Initial data processing steps included (i) lung segmentation from adjacent thoracic and chest wall structures, (ii) separation of right and left lungs and (iii) airway segmentation. Lung segmentation is performed using adaptive density-based morphological approach (described in appendix), and airway segmentation involved density thresholding. Right and left lungs were then segmented into upper, middle and lower zones (Figure A). The carina was used as a landmark to identify the lower boundary of the upper zone. The remaining two thirds of each lung were equally subdivided into the middle and lower zones for each lung. Total right and left lung volumes were calculated.

###### **Pulmonary vessel quantification**

To derived pulmonary vessel volume (PVV), pulmonary vessels were extracted from lung parenchyma using a multi-scale tubular structure enhancement filter. An algorithm determined likelihood that each voxel was connected to a dense tubular structure representing a blood vessel or blood vessel-related tubular structure (Figure B)^1^. This assessment excluded large vessels at the hilum of the lung.

The PVV was calculated as an absolute volume (cm^3^) and expressed as a relative percentage of total CALIPER-derived lung volume. Total and regional volumes were calculated. Regional volumes were expressed as upper, middle, and lower zone volume and calculated from sum of respective right and left lung zonal volumes. In cases that underwent a second CT, change in PVV is expressed as either (i) absolute change in PVV (CT2 – CT1), relative change in absolute PVV or absolute change in PVV% (CT2 PVV% - CT1 PVV%) or (ii) annualised change in each of these metrics relative to time interval between CT1 and CT2.

Pulmonary vessel volume (PVV) was also explored in a separate cohort of patients undergoing volumetric CT for routine follow up assessment of pre-existing lung nodules. This was to provide a meaningful comparison of PVV assessment in cases without fibrotic lung disease. All CT scans were non-contrast volumetric scans (0.625mm slice thickness at an interval of 0.625mm).

###### **Pattern evaluation**

After extracting the pulmonary vessels, parenchymal tissue type classification was applied to 15x15x15 voxel volumes of interest using texture analysis and computer-based algorithmic interpretation of volumetric histogram signature mapping features as previously described above (Figure C)^2^. CALIPER evaluation of CT data included classification and quantification of each volume of interest into one of six radiological parenchymal categories: normal lung, hyperlucent, ground glass opacity (GGO), Reticular, honeycombing. Volumes for each parenchymal feature were expressed as a relative percentage of CALIPER-derived (i) total lung volume or (ii) zonal lung volume. Total fibrosis extent represented the sum of GGO, reticular and honeycomb percentages.

In cases that underwent a second CT, change in fibrosis is expressed as (i) absolute change in total fibrosis extent (CT2 – CT1) and (ii) absolute change in total fibrosis extent per litre of lung volume (CT2 total fibrosis / litre – CT1 total fibrosis / litre). The latter is a standardised measure to account of difference in lung volume on serial scans. Progression of fibrosis can be observed with a simultaneous reduction in lung volume. As CALIPER reports parenchymal features as a percentage of total lung volume reported values could be influenced by this denominator. Therefore, accuracy in quantification of fibrosis progression is also influenced by this.

Annualised change in each of these metrics relative to time interval between CT1 and CT2.

The platform provided data output of DICOM series with a colour-coded overlay reflective of parenchymal abnormality and overall disease extent. This was used as a quality control check for each scan. A pdf file was also provided for derived metrics which were then collated into a database for analysis. See appendix.

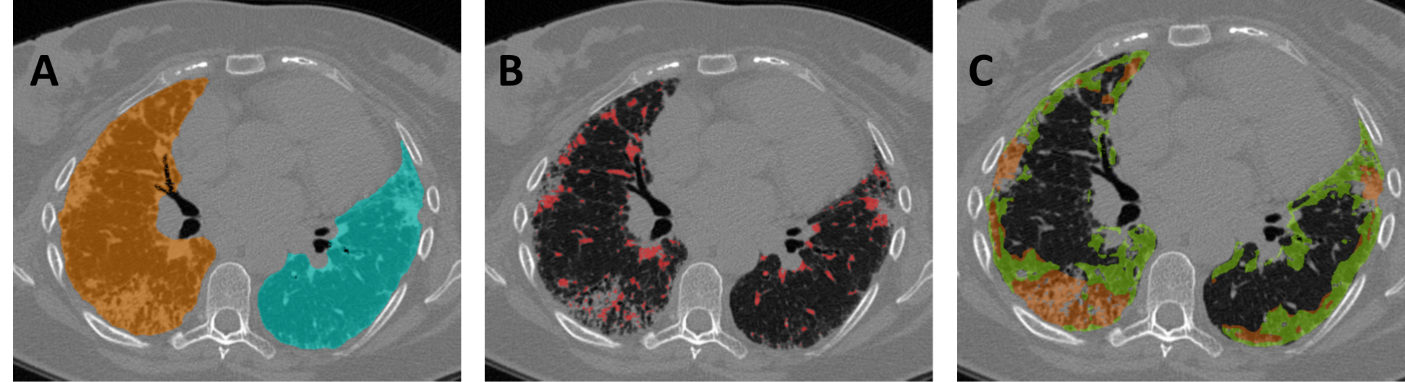

**Figure A-C** Axial slices of CALIPER LTA (A) lung labels, (B) pulmonary vessel labels, (C) lung texture analysis map.

|  | **MLR** | | **NLR** | | **SIRI** | |
| --- | --- | --- | --- | --- | --- | --- |
| **Baseline metrics** | **r** | **p** | **r** | **p** | **r** | **p** |
| **CALIPER scores** |  |  |  |  |  |  |
| TLF (%) | -0.046 | 0.558 | -0.044 | 0.491 | -0.011 | 0.881 |
| TLF (/litre) | -0.083 | 0.289 | 0.027 | 0.730 | 0.010 | 0.897 |
| PVV (cm^3^) | 0.085 | 0.283 | 0.039 | 0.622 | 0.067 | 0.397 |
| PVV (%) | 0.037 | 0.639 | 0.122 | 0.121 | 0.133 | 0.091 |
| **Lung function tests** |  |  |  |  |  |  |
| FVC% | -0.083 | 0.159 | -0.091 | 0.119 | -0.115 | **0.049** |
| TLCO% | -0.013 | 0.833 | -0.043 | 0.468 | -0.089 | 0.133 |
| CPI | 0.062 | 0.298 | 0.057 | 0.334 | 0.115 | 0.053 |

**Supplemental Table S1. Pearson (r) correlation matrix of derived leukocyte indexes and CALIPER parameters and lung function at baseline.** n=171 cases. PVV - pulmonary vessel volume, MLR - monocyte:lymphocyte ratio, NLR - neutrophil:lymphocyte ratio, SIRI; systemic inflammatory response index (neutrophil ×monocyte/lymphocyte

| **Disease progression** | **∆TLF <7.8%/Ltr**  **(Stable)** | **∆TLF >7.8%/Ltr**  **(Progression)** | **p Value** | **∆TLF <10%/Ltr**  **(Stable)** | **∆TLF >10%/Ltr**  **(Progression)** | **p Value** | **∆FVC <10%**  **(Stable)** | **∆FVC >10%**  **(Progression)** | **p Value** |
| --- | --- | --- | --- | --- | --- | --- | --- | --- | --- |
| **n** | 46 | 25 |  | 58 | 13 |  | 54 | 17 |  |
| Monocytes (x10^3^/μl) | 0.64 (0.55-0.80) | 0.75 (0.62-0.8) | 0.813 | 0.68 (0.55-0.82) | 0.66 (0.55-0.77) | 0.162 | 0.68 (0.57-0.88) | 0.63 (0.54-0.73) | 0.062 |
| Neutrophils (x10^3^/μl) | 4.66 (3.78-6.24) | 5.36 (5.26-8.08) | 0.845 | 4.84 (3.85-6.45) | 4.87 (3.86-5.66) | 0.043 | 5.03 (3.92-6.49) | 4.72 (3.86-5.87) | 0.568 |
| Lymphocytes (x10^3^/μl) | 1.66 (1.43-2.29) | 1.58 (1.27-2.14) | 0.511 | 1.66 (1.39-2.53) | 1.61 (1.47-2.01) | 0.568 | 1.76 (1.31-2.51) | 1.61 (1.45-2.08) | 0.639 |
| MLR | 0.37 (0.28-0.47) | 0.5 (0.33-0.69) | 0.599 | 0.38 (0.27-0.49) | 0.37 (0.32-0.58) | 0.158 | 0.42 (0.33-0.52) | 0.36 (0.27-0.42) | 0.156 |
| NLR | 2.60 (1.96-3.59) | 3.34 (2.55-3.83) | 0.827 | 2.60 (1.88-3.97) | 2.71 (2.35-3.34) | 0.091 | 2.66 (2.16-3.92) | 2.77 (2.24-3.58) | 0.924 |
| SIR | 1.71 (1.21-2.57) | 2.29 (1.72-3.79) | 0.489 | 1.77 (1.21-2.72) | 1.80 (1.5-3.09) | 0.064 | 2.08 (1.53-3.33) | 1.66 (1.27-2.29) | 0.115 |

Supplemental Table S2. Comparison of blood leukocytes in n=71 cases with stable or progressive disease at repeat CT scan

Data is expressed as medians (interquartile range). Difference between groups (stable vs progression) assessed using Mann-Whitney test. p<0.05 considered statistically significant.

| **Multivariate Cox regression** | **HR** | **95%CI** | **p Value** |
| --- | --- | --- | --- |

| **Model A: Increase in Fibrosis >7.8 % / litre** | | | | | | |
| --- | --- | --- | --- | --- | --- | --- |
| Model A1 | MLR | | 75.88 | | 3.8-1515 | **0.005** |
| Model A2 | NLR | | 1.44 | | 1.1-1.9 | **0.009** |
| Model A3 | SIRI | | 1.76 | | 1.19-2.6 | **0.005** |
| **Model B: Increase in Fibrosis >10.0 % / litre** | | | | | | |
| Model B1 | MLR | | 122.04 | | 3.76-3957.09 | **0.007** |
| Model B2 | NLR | | 1.57 | | 1.12-2.22 | **0.010** |
| Model B3 | SIRI | | 2.06 | | 1.25-3.38 | **0.004** |
| **Model C: Decline in FVC >10%** | | | | | | |
| Model C1 | | MLR | 4.12 | 0.3550.3 | | 0.258 |
| Model C2 | | NLR | 1.12 | 0.95-1.31 | | 0.174 |
| Model C5 | | SIRI | 1.20 | 0.93-1.54 | | 0.154 |

**Supplemental Table S3. Multivariate Cox regression for leukocyte variables for outcome of progression for dichotomised leukocyte levels and derived leukocyte indexes (MLR, NLR and SIRI)**

Hazard ratios in multivariate model for leukocytes and derived indexes modelled against outcomes of increase in fibrosis on follow on CT scan of TLF/litre $\geq$ 7.8 % /litre (Model A) and TLF/litre $\geq$ 10% /litre (Model B); or change in lung function of FVC decline >10% (Model C). Leukocyte derived indexes (MLR, NLR or SIRI) were modelled in separate models. All models adjusted for age, gender, % drop in CALIPER lung volume, PVV and LAA% (outcomes for these variables are not shown).

| **Outcome**  **Increase in TLF** $\geq$ **10% /litre between CT1 and CT2** | **HR (95% CI)** | **p** |
| --- | --- | --- |
| **Model D** |  |  |
| Monocytes (x10^3^/μl) | 3.08 (0.13-72.58) | 0.485 |
| Neutrophils (x10^3^/μl) | 2.23 (1.05-4.71) | **0.036** |
| Lymphocytes (x10^3^/μl) | 0.23 (0.03-1.57) | 0.132 |
| **Model E** |  |  |
| MLR | 76.15 (0.86-6770) | 0.058 |
| **Model F** |  |  |
| NLR | 1.59 (1.03-2.45) | **0.037** |
| **Model G** |  |  |
| SIRI | 2.08 (1.11-3.9) | **0.022** |

Supplemental Table S4. Cox Proportional hazard analysis for progression of fibrosis adjusted for antifibrotic use

Multivariate Cox proportional hazard analysis for outcomes of progression of fibrosis (increase in TLF $\geq$ 10% /litre)(Models with TLF $\geq$ 7.8% showed no significant correlation) . All models adjusted for age, gender, baseline fibrosis score and antifibrotic use at first CT scan. For multivariate model testing contribution of blood leukocytes against outcome these were all tested in combination [absolute monocyte, lymphocyte, and neutrophils] to explore interaction (Model D). For multivariate models exploring contribution of the leukocyte derived indexes [MLN, NLR or SIRI] these were tested individually and in absence of other leukocytes measurements or derived indexes (Models E-G). MLR; monocyte:lymphocyte ratio, NLR; neutrophil:lymphocyte ratio, SIRI; systemic inflammatory response index (neutrophil × monocyte/lymphocyte).

| **Multivariate Cox regression** | **HR (95% CI)** | **p Value** |
| --- | --- | --- |
| **Model H: Increase in TLF** ≥ **7.8%/litre between CT1 and CT2** | | |
| -Charlson comorbidity index | 0.67 (0.45-1.01) | 0.056 |
| -Male | 0.01 (0.00-0.20) | 0.013 |
| -Change in lung volume (%) | 0.84 (0.76-0.92) | <0.001 |
| -Total lung fibrosis (/litre) on 1st CT | 1.18 (1.08-1.29) | <0.001 |
| -Total PVV (%) | 1.04 (1.01-1.07) | 0.004 |
| -Low attenuation areas (%) | 1.09 (0.96-1.25) | 0.179 |
| -Monocyte (x10^3^/μl) | 2.13 (0.12-39.4) | 0.611 |
| -Neutrophil (x10^3^/μl) | 2.13 (1.06-4.3) | 0.035 |
| -Lymphocyte (x10^3^/μl) | 0.05 (0.01-0.39) | 0.004 |
| **Model I: Increase in TLF** ≥ **10%/litre between CT1 and CT2** | | |
| -Charlson comorbidity index | 0.93 (0.84-1.04) | 0.208 |
| -Male | 0.02 (0-0.46) | 0.016 |
| -Change in lung volume (%) | 0.82 (0.73-0.92) | <0.001 |
| -Total lung fibrosis (/litre) on 1st CT | 1.13 (1.03-1.25) | 0.013 |
| -Total PVV (%) | 1.04 (1-1.07) | 0.037 |
| -Low attenuation areas (%) | 1.1 (0.97-1.25) | 0.142 |
| -Monocyte (x10^3^/μl) | 2.37 (0.09-62.2) | 0.604 |
| -Neutrophil (x10^3^/μl) | 2.66 (1.35-5.25) | 0.005 |
| -Lymphocyte (x10^3^/μl) | 0.3 (0.07-1.24) | 0.096 |
| **Model J: Decline in FVC >10% between CT1 and CT2** | | |
| -Charlson comorbidity index | 1.06 (0.89-1.25) | 0.539 |
| -Male | 0.66 (0.17-2.52) | 0.539 |
| -%FVC baseline | 0.99 (0.97-1.01) | 0.243 |
| -Monocyte (x10^3^/μl) | 0.34 (0.04-3.14) | 0.341 |
| -Neutrophil (x10^3^/μl) | 0.93 (0.76-1.14) | 0.477 |
| -Lymphocyte (x10^3^/μl) | 0.82 (0.47-1.44) | 0.495 |

**Supplemental Table S5 Cox Proportional hazard analysis** **for progression of lung fibrosis**

Multivariate Cox proportional hazard analysis for outcomes of progression of fibrosis. Comorbidities adjusted for using Charlson comorbidity index.^1^ For multivariate model testing contribution of blood leukocytes against outcome these were all tested in combination.

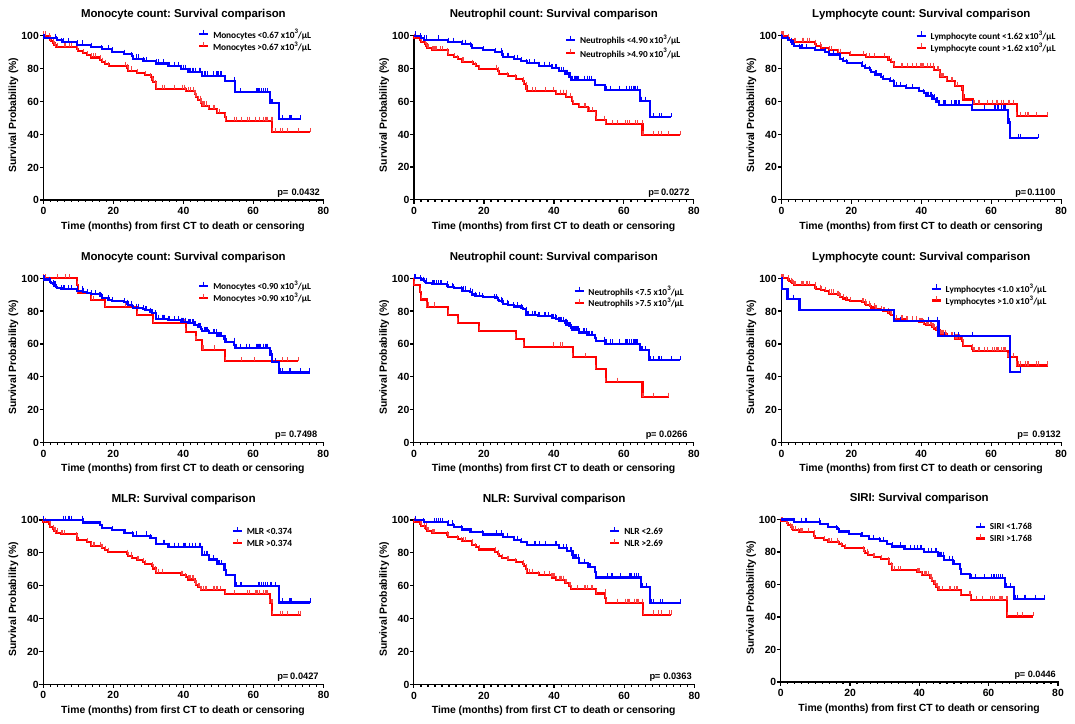

Supplemental Figure S1. Kaplan-Meier curves for time to mortality for monocyte, neutrophil, lymphocyte levels, MLR, NLR and SIRI, all at baseline. Values are dichotomised above and below median values (top panel, as shown in Figure 1), upper limit of normal reference range limits (middle panel) and for derived indexes of monocyte:lymphocyte ratio, neutrophil:lymphocyte ratio and systemic inflammatory response index (bottom panel)
